# Genome-Wide Meta-Analysis Validates a Role for *S1PR1* in Microtubule Targeting Agent-Induced Sensory Peripheral Neuropathy

**DOI:** 10.1101/2020.04.23.20076208

**Authors:** Katherina C. Chua, Chenling Xiong, Carol Ho, Taisei Mushiroda, Chen Jiang, Flora Mulkey, Dongbing Lai, Bryan P. Schneider, Sara R. Rashkin, John S. Witte, Paula N. Friedman, Mark J. Ratain, Howard L. McLeod, Hope S. Rugo, Lawrence N. Shulman, Michiaki Kubo, Kouros Owzar, Deanna L. Kroetz

**Author notes:** Corresponding Author: Deanna L. Kroetz, Ph.D., 1550 4^th^ Street RH584E, Box 2911, San Francisco, CA 94143-2911. Conflicts of interest: The authors declare no conflicts of interest.

## Abstract

Microtubule targeting agents (MTAs) are anticancer therapies commonly prescribed for breast cancer and other solid tumors. Sensory peripheral neuropathy (PN) is the major dose-limiting toxicity for MTAs and can limit clinical efficacy. The current pharmacogenomic study aimed to identify genetic variations that explain patient susceptibility and drive mechanisms underlying development of MTA-induced PN. A meta-analysis of genome-wide association studies (GWAS) from two clinical cohorts treated with MTAs (CALGB 40502 and CALGB 40101) was conducted using a Cox regression model with cumulative dose to first instance of grade 2 or higher PN. Summary statistics from a GWAS of European subjects (n = 469) in CALGB 40502 that estimated cause-specific risk of PN were meta-analyzed with those from a previously published GWAS of European ancestry (n = 855) from CALGB 40101 that estimated the risk of PN. Novel single nucleotide polymorphisms in an enhancer region downstream of sphingosine-1-phosphate receptor 1 (*S1PR1* encoding S1PR_1_; *e*.*g*., rs74497159, β_CALGB 40101_ per allele log hazard ratio (95% CI) = 0.591 (0.254 - 0.928), β_CALGB 40502_ per allele log hazard ratio (95% CI) = 0.693 (0.334 - 1.053); *P*_*META*_ = 3.62×10^−7^) were the most highly ranked associations based on P-values with risk of developing grade 2 and higher PN. *In silico* functional analysis identified multiple regulatory elements and potential enhancer activity for *S1PR1* within this genomic region. Inhibition of S1PR_1_ function in iPSC-derived human sensory neurons shows partial protection against paclitaxel-induced neurite damage. These pharmacogenetic findings further support ongoing clinical evaluations to target S1PR_1_ as a therapeutic strategy for prevention and/or treatment of MTA-induced neuropathy.

## Introduction

Microtubule-targeting agents (MTAs) such as taxanes and epothilones, are widely prescribed for treatment of various solid tumors. These mitotic inhibitors disrupt microtubule dynamics via stabilization of microtubules to primarily target and block rapidly dividing cells^1^, and are effective in treating primary and metastatic breast cancer. However, MTA therapy often causes significant dose-limiting toxicities, including sensory peripheral neuropathy. This nerve damage presents as distal axonal degeneration, and clinically manifests as numbness, tingling or painful sensations in a “glove and stocking” distribution^2,3^. Up to 50% of patients experience some degree of sensory peripheral neuropathy with as many as 30% reaching severe peripheral neuropathy (grade 3 or 4)^3,4^. These symptoms manifest as early as the first dosing cycle, and in severe cases, the symptoms persist for years after the last course of therapy^2,4^. Reported risk factors for drug-induced neuropathy include prior treatment with neurotoxic agents, frequency of chemotherapy dosing, high cumulative chemotherapy exposure, preexisting neuropathy and age^2–4^. However, these risk factors do not fully account for the observed incidence of chemotherapy-induced peripheral neuropathy (CIPN)^3^. There are currently no neuroprotective strategies that provide adequate clinical efficacy in preventing this toxicity and only duloxetine has been recommended for treatment of existing painful CIPN^3^.

Human genetic association studies have been used as tools to identify critical genes involved in the pathophysiology of CIPN. In genetic association studies, consideration of the highest-ranking single nucleotide polymorphisms (SNPs) with respect to P-value suggested a potential role for genes related to the regulation of neuron morphogenesis and neurodegeneration (*EPHA4/5/6, FGD4, FZD3, ARHGEF10, VAC14*) in the pathogenesis of taxane-induced peripheral neuropathy^5^. While most of these genetic association findings did not reach genome-wide significance, candidate gene analyses in independent populations support the associations with ephrin receptor genes, *FGD4*, and *ARHGEF10*^6^. The primary goal in the current study was to conduct a meta-analysis with two cohorts of breast cancer patients (CALGB 40502 and CALGB 40101) to extend these genomic findings and further elucidate this complex phenotype.

Cancer and Leukemia Group B (CALGB) 40502 was a phase III randomized three-arm study comparing nanoparticle albumin-bound (nab) paclitaxel or ixabepilone once per week to weekly paclitaxel as first-line therapy for patients with advanced breast cancer^7^; bevacizumab was administered in all arms of the study. CALGB 40101 was a phase III randomized trial with 2×2 factorial design to test the noninferiority of single agent paclitaxel with doxorubicin and cyclophosphamide (AC) and the superiority of six cycles of treatment over four cycles^8^. The most common grade 3 to 4 nonhematological toxicity in both studies was sensory peripheral neuropathy^7,8^. Findings from the pharmacogenomic association meta-analysis were functionally evaluated *in vitro* to probe the mechanistic basis of chemotherapy-induced sensory neuron damage.

## Materials and Methods

### Participants

All study participants were enrolled in either CALGB 40502 or CALGB 40101. Trial design and eligibility criteria for enrollment for each clinical trial have been previously described^7,8^. CALGB is now a part of the Alliance for Clinical Trials in Oncology. All subjects provided written informed consent for both the treatment and companion pharmacogenetic protocols that met state, federal, and institutional guidelines. Only subjects in CALGB 40101 receiving paclitaxel every two weeks were included in pharmacogenetic studies. Pharmacogenetic analysis was approved by Institutional Review Boards at the National Cancer Institute and the University of California San Francisco. Toxicity data was collected by the Alliance Statistics and Data Center, sample genotyping was performed at Riken Center for Genomic Medicine and statistical analysis was done at the University of California San Francisco. Replication was performed in samples from ECOG5103^9^ and the UK Biobank^10^.

### Genotype Data

Study characteristics of CALGB 40502 and CALGB 40101 are shown in Table 1. From the 799 patients randomized in CALGB 40502, 633 consented patients with DNA samples were genotyped using the Illumina HumanOmniExpressExome-8 BeadChip, interrogating 964,055 SNPs with coverage of common variants and additional exonic content. Genotyping data were filtered using a standard quality control (QC) pipeline (Figures S1-S3). Detailed sample and variant quality control are described in Supplementary Methods. Samples were filtered for low call rate or low genotyping performance, relatedness, and sex check as a measure for genotyping quality. Using principal components analysis and self-reported ethnicity, a total of 485 subjects with estimated European ancestry were selected for primary analysis to prevent population stratification bias. Genetic imputation was performed with 902,927 autosomal SNPs using the Michigan Imputation Server^11^. After post-imputation filtering, a total of 23,210,471 imputed SNPs remained and a total of 469 samples from CALGB 40502 with complete phenotypic data were used in the association analysis. All samples from CALGB 40101 used in the current meta-analysis were described in a prior GWAS^12^ and are briefly described in Supplementary Methods. A total of 14,676,818 post-QC SNPs and 855 samples from CALGB 40101 with complete phenotype information were used for the genome-wide association analysis. After filtering for SNPs with beta estimates available from the individual GWAS in each cohort, 6,030,476 SNPs were considered in the final results of the meta-analysis. Genotype and neuropathy data for European subjects in ECOG-5103 and for UK BioBank subjects have been previously described^9,10^.

**Table 1.** Study characteristics of pharmacogenetic cohorts in CALGB 40502 and CALGB 40101

|  | **CALGB 40502** | **CALGB 40101** |
| --- | --- | --- |
| Consented with DNA | 633 | 1029 |
| Post Quality Control | 628 | 1023 |
| White (Non-Hispanic/Non-Latino) | 478 (76%) | 855 (83%) |
| Discovery Cohort<sup>*</sup> | 469 | 855 |
| Disease Stage | Locally recurrent or metastatic breast cancer | Early-stage breast cancer with 0-3 positive axillary nodes |
| Drug therapy | Paclitaxel (90 mg/m<sup>2</sup>) or nab-paclitaxel (150 mg/m<sup>2</sup>) or ixabepilone (16 mg/m<sup>2</sup>) ± bevacizumab weekly for 3 of 4 weeks | Paclitaxel (175 mg/m<sup>2</sup>) biweekly for 4 or 6 cycles |
| Reported incidence of peripheral neuropathy<sup>**,†</sup> | 232 (49%) | 206 (24%) |
| Taxane-naïve<sup>**</sup> | 265 (57%) | 855 (100%) |
| Informative competing risks to developing peripheral neuropathy<sup>**</sup> | Yes | No |
| Reported incidence of diabetes<sup>**</sup> | 50 (12%) | 3 (<1%) |
| Genotyping array | Illumina HumanOmniExpressExome-8 BeadChip | Illumina HumanHap610-Quad Genotyping BeadChip |

<sup>\*</sup>Self-reported White (Non-Hispanic/Non-Latino) samples with complete phenotype information
<sup>\*\*</sup>Within discovery cohort
<sup>†</sup>Peripheral neuropathy is defined as NCI-CTCAE grade 2 or higher peripheral neuropathy event

### Phenotype Data

Adverse events, including chemotherapy-induced peripheral neuropathy, were graded according to the NCI Common Terminology Criteria for Adverse Events (NCI-CTCAE v3 in CALGB 40502; NCI-CTCAE v2 in CALGB 40101), defining the range of severity of neuropathy cases as grade 0-4. Since the incidence of the toxicity is dependent on cumulative drug exposure, sensory peripheral neuropathy was assessed with a dose-to-event phenotype. An MTA-induced sensory peripheral neuropathy event was defined as the cumulative MTA dose (mg/m^2^) to first instance of grade 2 or higher sensory peripheral neuropathy. In CALGB 40502, patients were treated until unacceptable toxicity, other complicating disease, alternative therapy, patient withdrawal, treatment completion, progression or death. Patients for whom no neuropathy event was reported were informatively censored at time of occurrence with one of the other competing risks. Patients in CALGB 40101 for whom no neuropathy event was reported were uninformatively censored at completion of treatment (*i*.*e*., four or six treatment cycles).

### Statistical Analysis for Genome-wide Analysis

Genome-wide association analyses were first individually completed for each CALGB cohort to obtain summary statistics for the discovery meta-analysis. In CALGB 40502, each SNP was tested for an association with cause-specific hazard of neuropathy event within the framework of a Cox model using the Wald statistic, stratifying for treatment arm and adjusting for age. In CALGB 40101, the association between genotypic variation for each SNP and hazard of neuropathy event was tested within the framework of a Cox model using the score statistic, assuming uninformative censoring. In each case, genotypic variation was inferred using imputed allele dosage for untyped SNPs and associations were tested assuming an additive genotype-phenotype effect. This analysis was conducted under the *R* statistical environment^13^ 3.3 with the GenABEL^14^, survival^15^, and cmprsk^16^ packages. Per SNP summary statistics were further used to conduct an inverse-variance weighted meta-analysis using the METAL software^17^. P-values and confidence interval estimates have not been adjusted to account for multiple testing. This discovery analysis used ranking of unadjusted P-values for prioritization for additional analyses. Details of the replication analyses are provided in the Supplementary Methods.

### In vitro neurotoxicity studies

Human induced pluripotent stem cells (iPSCs) were differentiated into mature sensory neurons (day 35+; iPSC-SNs) according to a published protocol^18^. iPSC-SNs were treated with paclitaxel (Sigma-Aldrich, cat. no. T7402; St. Louis, MO) with or without the S1PR_1_ modulators FTY720 (Cayman Chemical; cat. no. 11975) or W146 (Cayman Chemical, cat. no. 10009109; Ann Arbor, MI), and compared to those treated with 0.2% DMSO (Sigma-Aldrich, cat. no. D2650; St. Louis, MO) as a vehicle control. After 48 hours of drug treatment, sensory neurons were fixed in 4% paraformaldehyde for 15 min at room temperature. After washing with phosphate buffered saline (PBS), cells were permeabilized with 0.25% Triton-X (Sigma-Aldrich; Saint Louis, MO) in PBS for 10 min at room temperature. Cells were then blocked with the addition of 10% goat serum (Jackson ImmunoResearch Laboratories, Inc.; West Grove, PA) in 1% bovine serum albumin and 0.5% Tween-20 blocking solution for 1 hour. Fixed cells were incubated overnight at 4°C with anti-TUJ1 antibody (Covance, cat. no. MRB-435P; Princeton, NJ) to stain for neuron-specific class III beta-tubulin. Goat anti-rabbit secondary antibody (Life Technologies, cat. no. A-11008; Carlsbad, CA) was added in blocking buffer for 1 hour. After PBS washes, 4′,6-diamidino-2-phenylindole (DAPI) (ThermoFisher, cat. no. D1306; Waltham, MA) was added to stain for nuclei. Imaging was performed at 20X magnification using the IN Cell Analyzer 2000 (GE Healthcare Life Sciences; Pittsburgh, PA).

### Imaging Data Analysis

Workflow of the imaging analysis is shown in Figure S4. Nine raw images were generated from each well, representing a field-of-view of 15.95 mm^2^ (2048 × 2048 pixels; 47% of well area). These images were batch processed through an imaging processing software, MIPAR™, with a custom-built algorithm to analyze measurements for chemotherapy-induced neuronal damage. This algorithm generates optimized grayscale images by reducing overall noise and minimizing the amount of non-specific staining to identify and quantify the neurite networks within each field-of-view image. A subsequent segmentation algorithm was performed to estimate nuclei within each field-of-view image. After processing, each image or field-of-view yielded measurements of total neurite area and neuron count. Neurite area was defined by the total area of pixels captured within the identified TUJ1-stained network for each image. Cell counts were designated with DAPI-stained nuclei, rejecting DAPI-stained particles less than 50 pixels to exclude nonspecific DAPI staining. To get a global measurement for each well, total neurite area and total cell count were generated by summing measurements across the nine field-of-view images. Processed images included further in the analysis were required to pass quality control on a per-well basis to assess the quality of the neurons and images (Figure S4). For quality control purposes, all nine field-of-view images were stitched together using an in-house script to generate per-well images, using the Grid/Collection Stitching plugin in Fiji. Wells were included in quantitative analyses if the corresponding stitched image met the following criteria: 1) neurites covered ≥ 50% of the entire well, 2) no more than 3 field-of-view images (out of 9) were out-of-focus, and 3) a majority of the signal intensities captured were not from artifacts. Per-well quality control was performed manually by at least two investigators blinded to the treatments.

### Statistical Analysis for Image Analysis

For each experiment, all drug treatments were completed with 6-8 replicates on the same plate and raw neurite area measurements and cell counts from imaging data were averaged to obtain a mean total neurite area and cell count per condition. Mean total neurite areas and mean total cell counts were expressed as a ratio of drug-treated to vehicle-treated samples. Differences between relative ratios for the treatment groups were tested for significance by one-way ANOVA test using the function *aov* and subsequent post hoc multiple comparisons using unpaired, two-sided Student’s *t* test with the function *t*.*test* in *R*^13^ version 3.5.3. The effects of S1PR_1_ modulators on paclitaxel neurotoxicity were assessed by comparison to paclitaxel-treated cells. Experiments were repeated three times with independent neuron differentiations and the results represent the mean phenotype measurements from each differentiation. The reported P-values and confidence interval estimates have not been adjusted for multiple testing.

## Results

Differences in sample size, disease stage, drug therapy, and genotyping arrays between the two pharmacogenetic discovery cohorts are described in Table 1. Detailed patient characteristics of CALGB 40502, including distribution of age, race, ethnicity, prior taxane status, and tumor subtype are listed in Table S1. Patient characteristics of the 615 subjects with complete genotype and phenotype information were similar to the 799 subjects randomized to the three-arm treatment in CALGB 40502. Characteristics of the CALGB 40101 subjects were previously summarized^12^, and are briefly described in Table S2. The main non-hematological toxicity in CALGB 40502 and CALGB 40101 was sensory peripheral neuropathy with reported event rates of grade 2 or higher of 49% and 24%, respectively (Table 1). Within CALGB 40502, a similar cumulative incidence of peripheral neuropathy was observed regardless of treatment arm (Figure 1; Figure S5), where risk of developing peripheral neuropathy was a function of cumulative chemotherapy dose. The main competing risk for developing peripheral neuropathy in the nab-paclitaxel arm was disease progression or death, while other competing risks, such as other adverse events, other complicating disease, alternative therapy, patient refusal, and treatment completion were more likely to lead to censoring for the peripheral neuropathy phenotype in the paclitaxel and ixabepilone arms. Within CALGB 40101, the cumulative incidence of paclitaxel-induced peripheral neuropathy has been previously reported^12^. Patients were uninformatively censored at four or six cycles if no peripheral neuropathy event was reported and therefore, no competing risks for developing peripheral neuropathy were considered.

**Figure 1.**
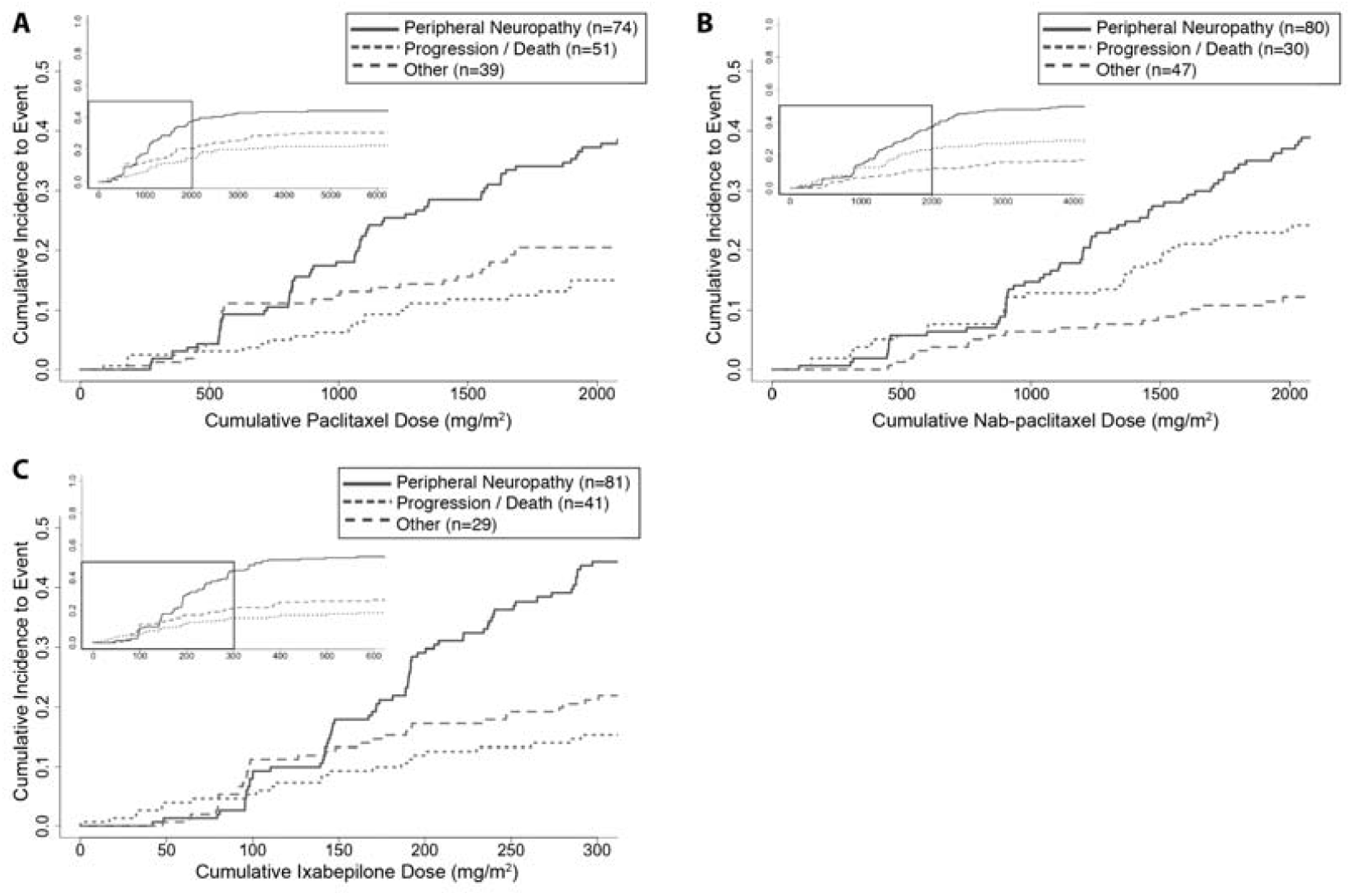
Cumulative incidence plot of chemotherapy-induced peripheral neuropathy (A: paclitaxel; B: nab-paclitaxel; C: ixabepilone) and informative competing events as a function of cumulative dose (mg/m^2^) to event for all subjects in the pharmacogenetics discovery cohort of CALGB 40502. Top left insert displays the entire range of cumulative doses, where the boxed area denotes where ∼75% of the data lies (Figure S5) and is the dose range represented in the larger plot.

The meta-analysis of the SNP associations from CALGB 40502 and CALGB 40101 was filtered for SNPs with a minor allele frequency of ≥ 5% in at least one of the two cohorts. None of the resulting SNPs reached genome-wide significance, although 18 linkage disequilibrium-pruned SNPs (r^2^ ≥ 0.7) had *P* < 10^−5^ (Table 2; Figures S6-S7). The 18 SNPs with the lowest P-values were filtered for further support of association from SNPs in high linkage disequilibrium with the identified SNP and expression in human dorsal root ganglion (DRG)^19^. SNPs in genomic regions annotated to *C9orf106, SLITRK1, KLHL1, LOC100129716*, and *SEPT5* had limited linkage support from visual inspection of LocusZoom^20^ plots and 11 SNPs were annotated to genes that are not detected in human DRG^19^ (*ZFPM2* (three independent SNPs), *C9orf106, KLHL1, SUGCT* (two independent SNPs), *ZBBX, LOC100129716, ADGRB3* and *CNGB1*). Based on the expression and linkage support filtering, additional analyses were only considered for the genomic regions around five SNPs (rs74497159/*S1PR1*, rs10771973/*FGD4*, rs11076190/*CX3CL1*, rs9623812/*SCUBE1*, rs2060717/*CALU*; Figures S8-12). Four of the remaining five SNPs were associated with increased risk of MTA-induced peripheral neuropathy, while a single SNP was protective. Cumulative incidence plots for MTA-induced peripheral neuropathy stratified by each of the five SNPs of interest are shown in Figure 2 and Figures S13-S16. Interestingly, the association of rs11076190 with peripheral neuropathy is driven by the paclitaxel-treated patients. None of these top five SNP associations were replicated in ECOG-5103 (Table S3) or the UK BioBank (Table S4).

**Table 2.**
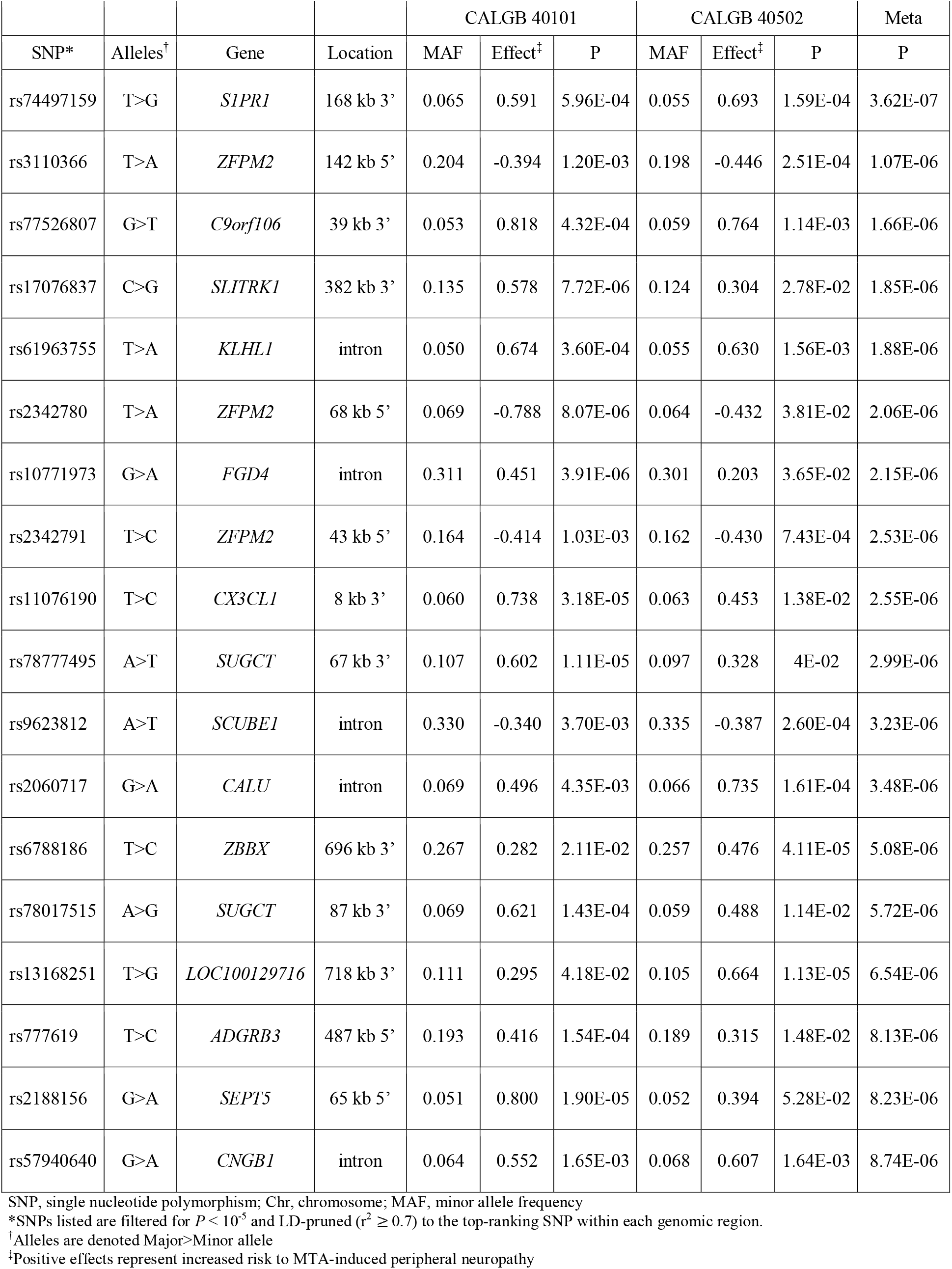
Top ranking SNPs for meta-analysis using cumulative dose to first instance of Grade 2 or higher peripheral neuropathy event.

**Figure 2.**
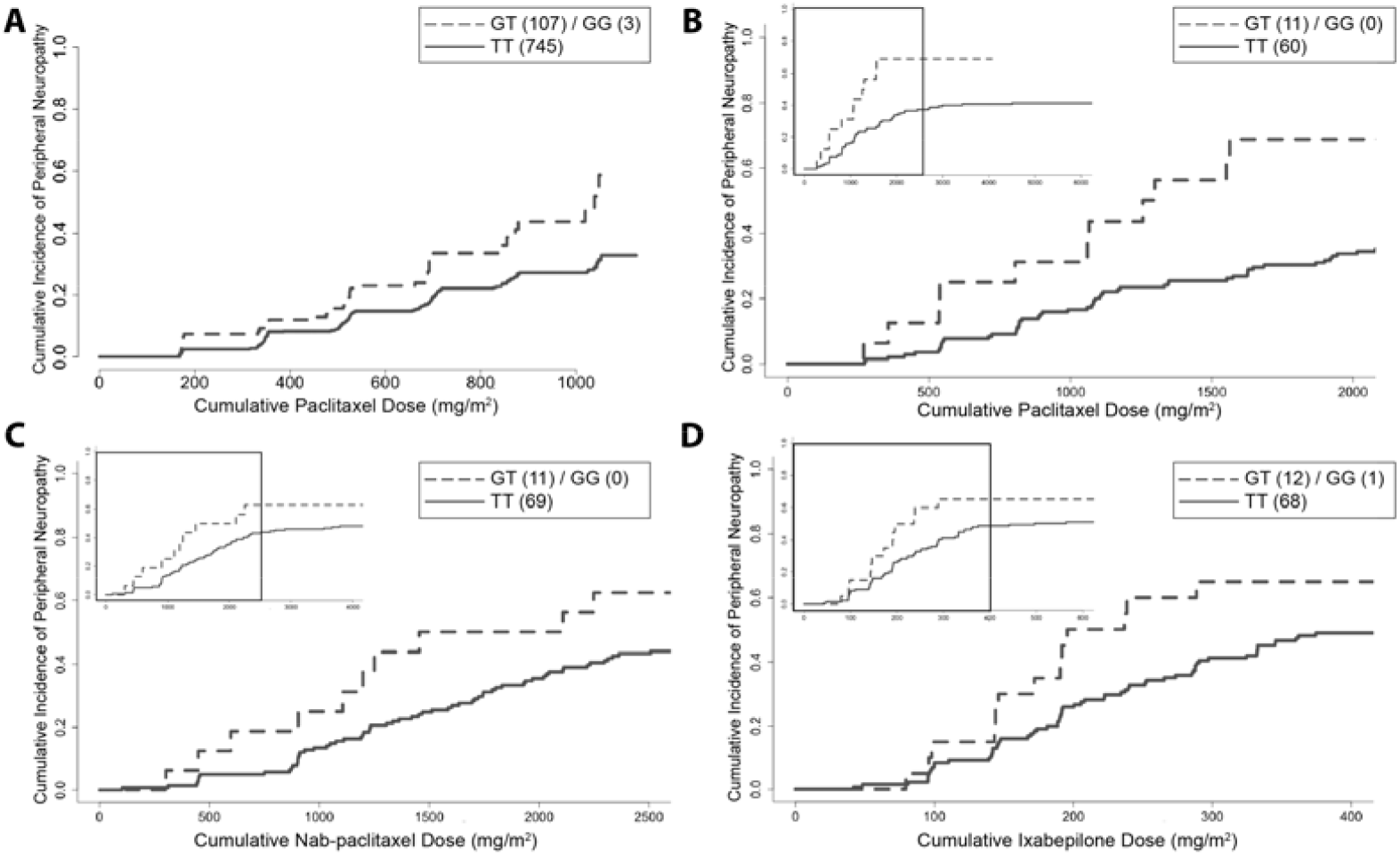
Cumulative incidence plot for chemotherapy-induced peripheral neuropathy stratified by rs74497159/*S1PR1* genotype in CALGB 40101 (A, paclitaxel) and CALGB 40502 (B, paclitaxel; C, nab-paclitaxel; D, ixabepilone). Top left insert displays the entire range of cumulative doses, where the boxed area denotes where ∼75% of the data lies (Figure S5) and is the dose range represented in the larger plot. The number of individuals with each genotype group is noted in parentheses. While the heterozygous and homozygous alternate genotype groups are grouped due to the limited number of observations, the allele risks represented in this cumulative incidence plot are estimated using an additive genetic model. The allele risk for peripheral neuropathy events without other competing events are shown in the plots for CALGB 40502 (B-D).

Bioinformatic analysis of the genomic regions surrounding the five SNPs chosen for further analysis was carried out to understand the potential functional effects of genetic variation on gene expression and function; the results from these *in silico* analyses are summarized in Tables 3 and S5. Examination of ENCODE data tracks (UCSC Genome Browser; https://genome.ucsc.edu) identified histone acetylation and methylation marks, DNase peaks, multiple transcription factor binding sites, and predicted functional activity from genome segmentation algorithms within the genomic regions surrounding rs74497159/*S1PR1*, rs10771973/*FGD4*, rs11076190/*CX3CL1*, rs9623812/*SCUBE1* and rs2060717/*CALU* (Figures S17-21). Further evidence that these SNPs are located in transcriptionally active regions includes classification as super-enhancers that interact with multiple genes. In some cases, the predicted enhancer region is expected to directly interact with the annotated gene to control its expression (Table 3). For example, a SNP annotated to *S1PR1* (rs74497159) is located downstream of the 3’ end of this gene in a super-enhancer region that interacts with *SIPR1*. Similarly, the intronic SNP of *FGD4* (rs10771973) lies adjacent to the last exon within a predicted transcriptional transition or elongation region and interacts directly with *FGD4*.

**Table 3.**
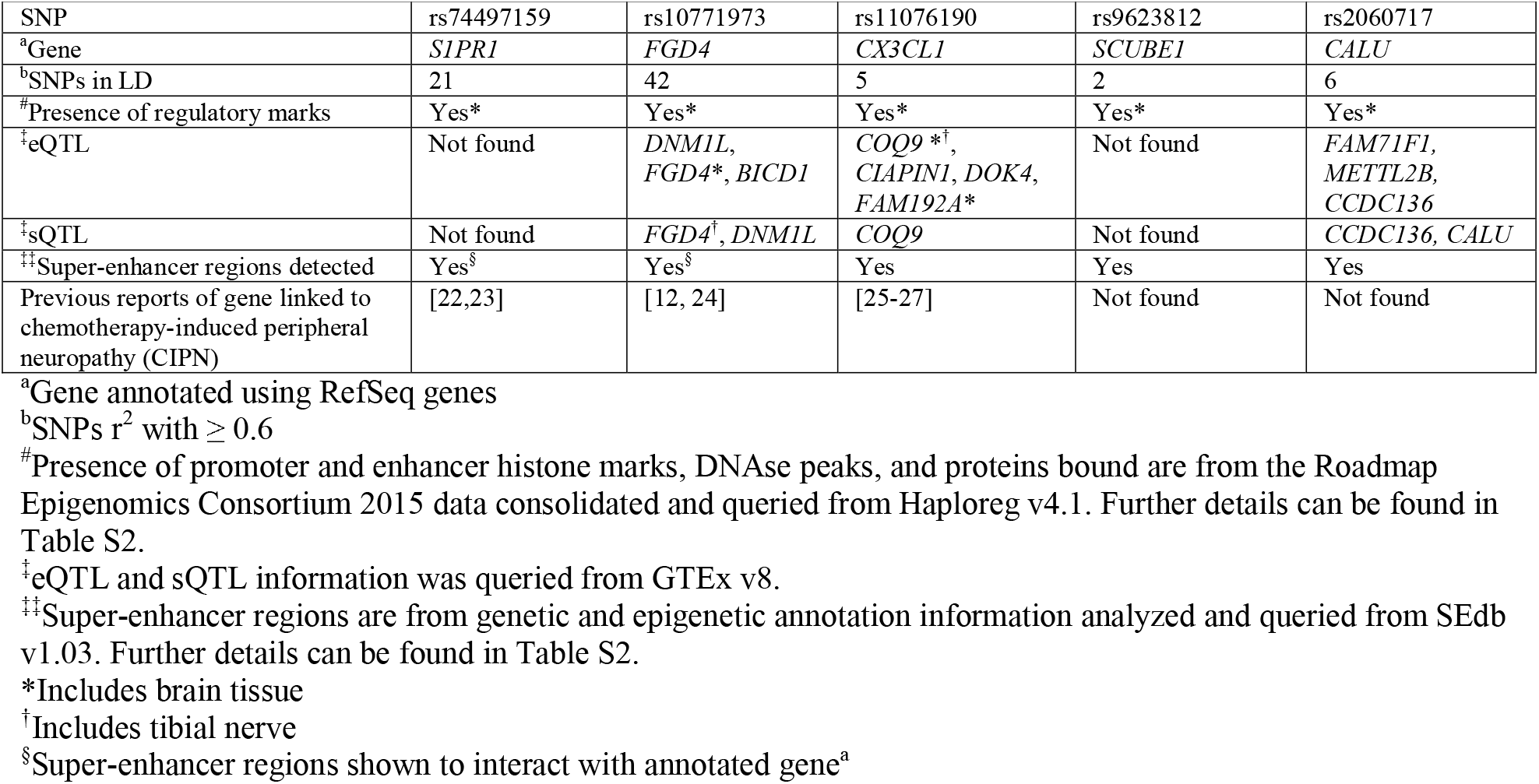
Summary of *in silico* functional analysis on LD (r^2^ ≥ 0.6) block of rs74497159, rs10771973, rs11076190, rs9623812, and rs2060717.

Intronic SNP rs10771973 is significantly associated with splicing quantitative trait loci in tibial nerve tissue (*P* = 1.2E-11, GTEx^21^), suggesting that this variant may regulate alternative splicing of pre-mRNA levels and affect the overall *FGD4* gene expression. rs11076190 is located in a genomic cluster with several chemokines (Figure S10) and is annotated within a FOXA1 transcription factor binding site linked to *CX3CL1* (Open Regulatory Annotation, ORegAnno, track; Figure S19). SNPs within linkage disequilibrium with rs10771973 (*FGD4*), rs11076190 (*CX3CL1*) and rs2060717 (*CALU*) are each associated with expression quantitative trait loci and/or splicing quantitative trait loci, indicating potential relevance for regulation of gene expression.

While the bioinformatic analysis highlights the potential functional activity for five SNPs of interest, the annotated genes from three of the five SNPs (rs74497159/*SIPR1*, rs10771973/*FGD4*, and rs11076190/*CX3CL1*) are linked to CIPN (Table 3). For functional validation, we focused on the gene annotated to the genomic region with the highest ranking based on P-value for association to MTA-induced peripheral neuropathy from our genome-wide meta-analysis, *S1PR1*. Since rs74497159 is annotated to the *S1PR1* gene and this SNP and others in linkage disequilibrium are in a super-enhancer region that controls the expression of *S1PR1*, the effect of modulation of S1PR_1_ functional activity was tested in human iPSC-SNs.

The human iPSC-SNs were generated following a published protocol and yield neurons expressing expected sensory neuron markers^18^. Based on paclitaxel dose-response studies, paclitaxel treatment (1 µM) for 48 hours in the iPSC-SNs had no significant effect on caspase-3/7 activity and decreased cellular ATP levels by <30%, indicating limited cytotoxicity under the conditions used (Figure S22). Drug-induced neurotoxicity is phenotypically characterized by a distinctive loss of neurites and a reduction in neurite network complexity without a decrease in total cell count (Figure 3; Figures S23-S24), which was quantified by total neurite area stained for βIII-tubulin and number of DAPI-stained nuclei. There was no significant effect of any treatment on cell numbers, consistent with limited cytotoxicity from paclitaxel and other chemicals. Treatment with 1 μM paclitaxel alone resulted in more than 50% decrease in neurite staining (49-64% reduction, *P* < 0.01; Figure 3) compared to vehicle-treated sensory neurons, demonstrating paclitaxel-induced damage to the overall neurite networks. Treatment with 1 μM S1PR functional antagonist FTY720 (0.3-29% reduction) and 1 μM S1PR_1_ antagonist W146 (0.6-14% reduction) had little to no effect on neurite area (Figure 3 and Figure S23). Combined treatment of the iPSC-SNs with paclitaxel and FTY720 resulted in partial protection against paclitaxel-induced neuronal damage (33-55% increase in neurite area relative to paclitaxel treatment, *P* < 0.05) (Figure 3 and Figure S23). The combination of paclitaxel and W146 had minimal effect on the paclitaxel-induced loss of neurite area (4-21% increase in neurite area relative to paclitaxel treatment; Figure 3 and Figure S23).

**Figure 3.**
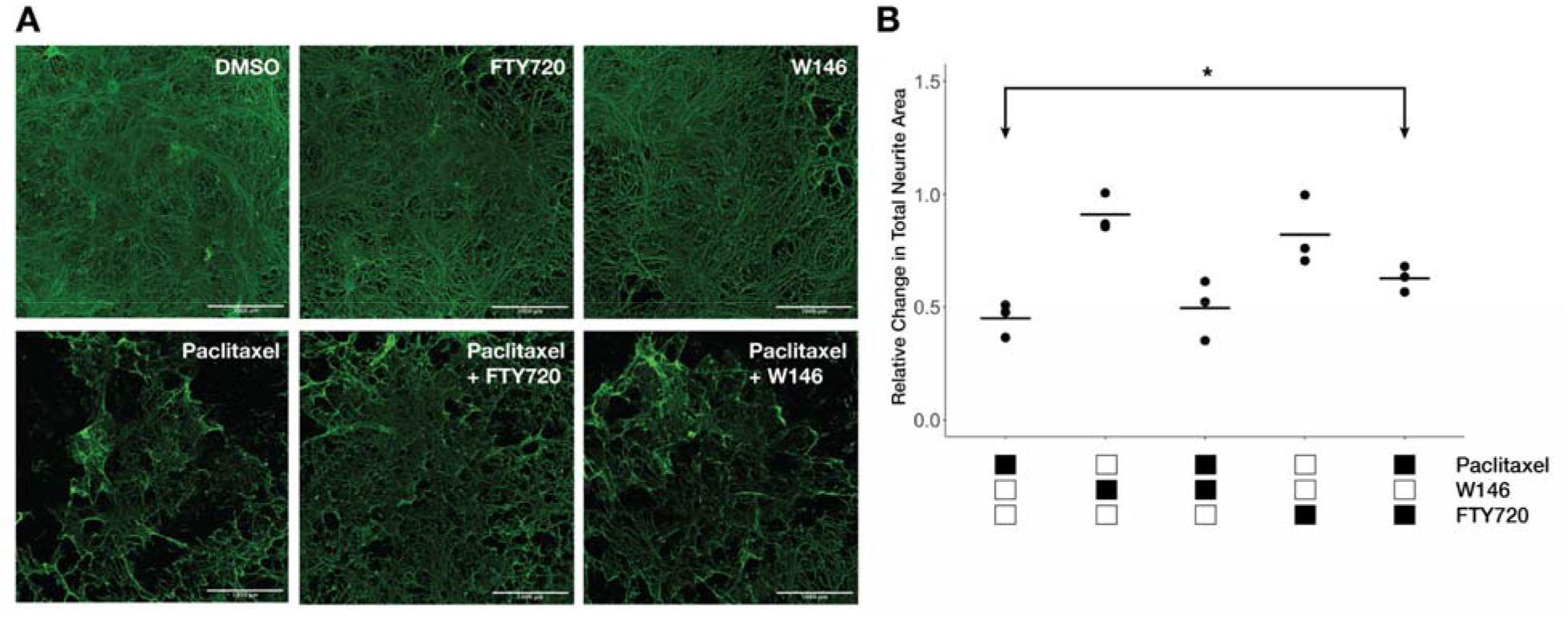
Inhibition of S1PR signaling attenuates paclitaxel-induced neuronal damage. (A) Representative per-well images of differentiated sensory neurons (D35+) derived from induced pluripotent stem cells used to investigate S1PR signaling in paclitaxel-induced neuronal damage. Differentiated neurons were treated with 1 µM paclitaxel for 48 hours in the absence and presence of a S1PR_1_ modulator (1 µM W146 or 1 µM FTY720), which both target and reduce signaling activity of S1PR_1_. W146 competitively binds and inhibits S1PR_1_ while the functional antagonist FTY720 reduces S1PR_1_ function by binding, internalizing and degrading the receptor. The cells are stained for βIII tubulin and staining was quantified as total neurite area. All images shown are from a single experiment. Scale bar indicates 1 mm. (B) Quantification of mean total neurite area from βIII tubulin staining after drug treatments in three independent differentiations. Each data point represents the mean measurement of 6-8 replicates from a single independent differentiation and expressed relative to vehicle controls. The coefficient of variation in vehicle-treated neurites ranges from 11-24%. Raw values used to calculate the means are shown in Figure S23. Relative mean neurite areas were tested for differences across treatments by one-way ANOVA (*P* = 6E-05) with post-hoc comparisons using unpaired, two-sided Student’s *t* test (\**P* < 0.05).

## Discussion

We identified multiple SNPs that implicate genes of potential relevance to MTA-induced sensory peripheral neuropathy, even though no SNP associations achieved genome-wide significance. Because the study focuses on sensory neuronal mechanisms involved in MTA-induced PN, five independent SNP associations with linkage disequilibrium support for association and whose nearest gene is expressed in human DRG^19^ were prioritized for *in silico* functional analysis to determine if the SNP lies in a potential regulatory genomic region. Three of the five SNPs (rs74497159, rs10771973, rs11076190) had the strongest *in silico* evidence for predicted functional activity and previous reports linking their annotated genes (*S1PR1*^22,23^, *FGD4*^12,24^ and *CX3CL1*^25–27^) to chemotherapy-induced neurotoxicity. Importantly, S1PR_1_ function was linked to neurotoxicity of paclitaxel in sensory neurons *in vitro*.

Among the three genomic regions identified from the primary meta-analysis, the highest-ranking association based on P-values revealed the genomic region in chromosome 1 annotated to *S1PR1*, a gene that encodes for sphingosine-1-phosphate receptor 1 (S1PR_1_). S1PR_1_ is a member of a G-coupled receptor family that is activated by its signaling ligand sphingosine-1 phosphate (S1P)^28^. In peripheral neurons, the S1P-to-S1PR_1_ axis has been associated with increased neuronal excitability^29^, reduction in neuronal growth through Rho GTPase signaling^30^, and increased hyperalgesia^31^ and other pain-like behaviors^32,33^. The SNP with the strongest association with MTA-induced peripheral neuropathy (rs74497159) is located in a genomic region experimentally defined as an enhancer with evidence for direct chromatin interactions with *S1PR1* and numerous eQTLs for *S1PR1*. Collectively, these bioinformatic data provide strong evidence that the identified genomic enhancer regions can regulate *S1PR1* expression, although additional studies will be needed to extend these data to regulation of *S1PR1* expression in peripheral sensory neurons. Considering these genomic findings and recent evidence highlighting S1PR_1_ as a drug target for prevention of chemotherapy-induced neuropathic pain *in vivo*^22,23^, functional studies investigated whether modulating S1PR_1_ function in sensory neurons would protect against MTA-induced damage.

Functional studies were performed using a human iPSC-derived sensory neuron model that displays paclitaxel-induced neurodegeneration. Two S1PR_1_ modulators with distinct mechanisms of inhibition were evaluated in these studies. Fingolimod (FTY720) binds, internalizes and degrades S1PR_1_ as a functional antagonist, and W146 competitively inhibits the receptor. FTY720 attenuated paclitaxel-induced neurotoxicity with similar effect sizes as previous functional validation studies in cellular models of chemotherapy-induced neuronal damage^34,35^. Additionally, functional antagonists of S1PR_1_ have consistently alleviated CIPN and other pain-like symptoms *in vitro* and *in vivo*^22,23,36,37^, and have led to ongoing phase I clinical trials investigating the use of fingolimod to prevent and treat chemotherapy-induced neuropathy (NCT03943498, NCT03941743). While treatment with the S1PR_1_ inhibitor W146 has also been shown to mitigate paclitaxel-induced neuropathic pain^22^ and S1P-induced hypersensitivity and thermal sensitivity *in vivo*^22,31^, minimal effects were observed with W146 treatment in the current *in vitro* studies. It is possible that a decrease in S1PR_1_ expression from internalization and degradation^38^ is essential to mitigate the effects of paclitaxel in peripheral neurons and that only blocking S1PR_1_ function with W146^39^ does not have the same pronounced protective effect. Interestingly, activation of S1PR_3_ may also be involved in sensory neurite retraction, nociceptor excitability, and pain-like symptoms^29,36,40^. FTY720 has also been shown to bind to S1PR_3_ and other receptors^38^, although its functional activity is largely attributed to S1PR_1_ binding. While expression of both S1PR_1_ and S1PR_3_ are known in primary DRG^41^ and iPSC-derived sensory neurons used in these studies (Figure S25), the exact role of these receptors in sensory peripheral neuropathy is not yet clear. These functional studies are the first step in understanding the role of S1PR_1_ signaling in peripheral sensory nociceptors under chemotherapy exposure and further investigations are needed to fully elucidate the role of sphingosine signaling in MTA-induced neuropathy.

The intronic SNP rs10771973 of *FGD4* was in the top SNPs from the meta-analysis and was the only variant identified in the previous GWAS in CALGB 40101 that remained in this meta-analysis^12^. Of note, *FGD4* is a critical gene for peripheral nerve development and a known causal gene of Charcot-Marie-Tooth subtype 4H, which is characterized by distal muscle weakness, severe foot deformities, sensory weakness or loss, and gait instability^42^. With this prior knowledge, it is conceivable that those harboring common genetic variation in *FGD4* may be more susceptible to peripheral neurotoxicity upon drug exposure. Importantly, in another genetic association study in an independent population, rs10771973 was linked with increased risk of paclitaxel dose reductions^24^. Further work in peripheral sensory neurons would reveal the exact mechanism of *FGD4* function in development of chemotherapy-induced neuropathy.

This genome-wide association study has also highlighted SNPs in a genomic region near *CX3CL1* and other chemokine genes. The top SNP (rs11076190) in this region lies just downstream of *CX3CL1*, encoding for a small chemokine ligand fractalkine (CX_3_CL_1_) that exclusively binds to CX_3_CR_1_ on lymphocytes. Paclitaxel treatment in preclinical models has been shown to increase levels of monocyte infiltration and inflammatory macrophage activation within peripheral nerves through CX_3_CL_1_-CX_3_CR_1_ crosstalk, releasing pro-inflammatory cytokines (TNF-α, IL-1β) and initiating peripheral neuropathic pain^43^. Additionally, previous work suggests that increased recruitment of transcription factors (i.e., NF-κB) to the *CX3CL1* promoter region is heightened in CIPN *in vivo* models^26^. Two additional genomic regions warrant investigation of potential involvement in CIPN. rs2060717 lies in a highly predicted promoter site within an intronic region of *CALU*, a gene that encodes for calcium-binding protein calumenin, which localizes to the endoplasmic reticulum with potential roles in early neuronal development^44^. The rs9623812 SNP lies in a potential enhancer site within an intronic region of *SCUBE1*, encoding for a cell surface glycoprotein that is secreted during brain injury^45^. Further studies are needed to understand the functions of these genes in sensory neurons and involvement in MTA-induced PN.

While this pharmacogenetic study using human genomic and cellular data has identified potential genes that have a translatable relevance to the CIPN phenotype, there are several limitations. Although a total of 1,324 samples from the discovery cohorts was used in the meta-analysis, the pharmacogenetic study presented is still insufficiently powered and increasing sample size may provide a more robust analysis. The main limitation is genetic validation of our top-ranking SNPs. None of the top-ranking SNPs (*P* < 10^−5^) from our meta-analysis were replicated in the taxane-treated ECOG-5103 European cohort or in the UK BioBank. In the case of ECOG-5103, paclitaxel treatment is different from both CALGB 40502 and CALGB 40101 and this sample was also limited by cohort size. While the UK BioBank is proving to be a useful resource for replication of genetic associations with common phenotypes, the largest number of drug-induced polyneuropathy cases identified was 122 subjects with ICD-9 or ICD-10 codes designated to “polyneuropathy due to drugs” and “polyneuropathy due to toxic agents”. While extending the analysis to subjects with other neuropathy types can increase sample size, this also introduces more heterogeneity into the phenotype. The other main limitation is the use of NCI-CTCAE grading for phenotyping peripheral neuropathy events, which has been shown to underestimate the progression of neuropathy symptoms and embody inconsistency in scale interpretation^46^. It is likely that phenotyping a combination of patient-reported and physician-reported outcomes will yield more comprehensive information. Additionally, while the genomic SNP region annotated to *S1PR1* lies within an active super-enhancer region supported by experimentally validated gene-enhancer interaction data from various cells/tissues, further validation of such interactions within sensory neurons are necessary to understand the direct SNP effect on enhancer activity and thus S1PR_1_ function. Lastly, our functional studies were limited to studying effects of S1PR_1_ in sensory neurons and did not investigate potential cross-talk between neurons and other cell types in the periphery. As CX_3_CL_1_ and S1PR_1_ both play established roles in peripheral lymphocytes, it is possible that their effects on paclitaxel-induced damage to peripheral nerves are initiated by external cues from other cell types. This phenomenon is true with *FGD4*, where the interplay of the frabin (*FGD4*)-Cdc42 Rho GTPase axis in Schwann cells causes peripheral nerve demyelination in CMT4H^47^.

In conclusion, this genome-wide association meta-analysis has identified potential genetic markers of MTA-induced peripheral neuropathy. This pharmacogenetic study validates the importance of S1PR_1_ receptor signaling from a genome-wide discovery analysis using clinical samples and functional validation using human iPSC-derived sensory neurons. Of note, S1PR_1_ signaling functionally intersects between both Rho-GTPase signaling and neuroinflammation, which have been well-documented to play roles in MTA-induced peripheral neuropathy. These findings provide further evidence for the role of sphingosine-1-phosphate signaling as a novel and exciting strategy for prevention and/or treatment of chemotherapy-induced neuropathy.

## Study Highlights

### What is the current knowledge on the topic?

Microtubule targeting-agent induced sensory peripheral neuropathy is a dose-limiting toxicity that can impact clinical benefit and significantly hinder quality of life. While some key genes involved in development of MTA-induced PN have been discovered from previous human genetic association studies, there is still limited understanding of the mechanisms underlying this common toxicity.

### What question did this study address?

This genome-wide meta-analysis was motivated to extend known genomic findings and discover novel targets that further elucidate the biology involved in MTA-induced PN.

### What does this study add to our knowledge?

Our pharmacogenetic approach using genome-wide data from clinical samples and functional studies in human sensory neurons has validated sphingosine-1-phosphate receptor signaling as a potential molecular driver of developing MTA-induced PN.

### How might this change clinical pharmacology or translational science?

The validation of sphingosine-1-phosphate receptor signaling in this study provides pharmacogenetic evidence to support current clinical investigations targeting this pathway for prevention or treatment of chemotherapy-induced peripheral neuropathy.

## Data Availability

Genotype and phenotype data into NCBI dbGAP submission is in progress.

## ClinicalTrials.gov Identifier

NCT00785291 (CALGB 40502) and NCT00041119 (CALGB 40101)

## Acknowledgements

The authors would like to thank all the patients and clinicians who took part in the study. The work presented in this study was completed as part of the dissertation from K.C.C.

## Author Contributions

K.C.C., L.N.S, M.J.R, H.L.M, K.O., and D.L.K wrote the manuscript. K.C.C, K.O., and D.L.K. designed the research. K.C.C., C.X., C.H., T.M., C.J., F.M., D.L., B.P.S., S.R.R., J.S.W., P.N.F, H.S.R, L.N.S, and M.K. performed the research. K.C.C., C.H., C.J., D.L., B.P.S., S.R.R., J.S.W., K.O., and D.L.K analyzed data.

